# Intra-host evolution during relapsing parvovirus B19 infection in immunocompromised patients

**DOI:** 10.1101/2024.10.04.24314882

**Authors:** Anne Russcher, Yassene Mohammed, Margriet E.M. Kraakman, Xavier Chow, Eric C.J. Claas, Manfred Wuhrer, Ann C.T.M. Vossen, Aloys C.M. Kroes, Jutte J.C. de Vries

## Abstract

**Introduction:** Parvovirus B19 (B19V) can cause severe relapsing episodes of anemia in immunocompromised individuals, which are commonly treated with intravenous immunoglobulins (IVIG). Few data is available on B19V intra-host evolution and the role of humoral immune selection. Here, we report the dynamics of genomic mutations and subsequent protein changes during relapsing infection.

**Methods:** Longitudinal plasma samples from immunocompromised patients with relapsing B19V infection in the period 2011-2019 were analyzed using whole genome sequencing to evaluate intra-host evolution. The impact of mutations on the 3D viral protein structure was predicted by deep neural network modeling.

**Results:** Of the three immunocompromised patients with relapsing infections for 3 to 9 months, one patient developed two consecutive nonsynonymous mutations in the VP1/2 region: T372S/T145S, and Q422L/Q195L. The first mutation was detected in multiple B19V IgG seropositive follow-up samples, and resolved after IgG seroreversion. Computational prediction of the VP1 3D structure of this mutant showed a conformational change in proximity of the antibody binding domain. No conformational changes were predicted for the other mutations detected.

**Discussion:** Analysis of relapsing B19V infections showed mutational changes occurring over time. Resulting amino acid changes were predicted to lead to a conformational capsid protein change in an IgG-seropositive patient. The impact of humoral response and IVIG treatment on B19V infections should be further investigated to understand viral evolution and potential immune escape.

## Introduction

Parvovirus B19 (B19V) is a 5.6 kb, single-stranded DNA virus. The viral genome codes for three major proteins: the viral capsid consists of a VP1 and VP2 protein, which share part of their coding sequences, while the non-structural (NS) protein is involved in replication and cellular processes [1]. Most infections occur in childhood and their presentation in children with fever and rash is known as erythema infectiosum or ‘fifth disease’, a mild self-limiting disease. However, due to the unique tropism of B19V for erythroid progenitor cells, B19V may cause severe anemia in certain susceptible populations. In severely immunocompromised individuals such as stem-cell or solid organ transplantation patients, this may lead to the clinical syndrome of ‘pure red cell aplasia’ (PRCA). PRCA can be treated by administering of intravenous immunoglobulins (IVIG), which usually leads to a reduction of viral load and alleviation of symptoms [2]. Because this effect of IVIG disappears with decreasing IVIG titer, a pattern of recurrent infections may occur, each episode managed by IVIG until the patient’s own immunity is restored [3].

In immunocompromised individuals, the inability to clear common self-limiting infections can lead to long term persistence of viruses, creating a reservoir in which mutants may arise. Immune escape mutants have been observed during prolonged infections with a variety of viral pathogens, such as SARS-CoV-2 and influenza virus, as well as latent DNA viruses including cytomegalovirus (CMV), during antiviral treatment [4-7]. Due to the relapsing nature of B19V in immunocompromised individuals, we hypothesize that new viral variants may also emerge in B19V infections over time.

Previous studies showed some genetic drift in prolonged B19V infection in patients treated with IVIG; small numbers of point mutations have been reported in single case reports or small case series [8-11]. These studies used conventional Sanger sequencing for analysis and mostly looked at partial genomes, focusing on capsid proteins. Over the past decade, the rapid development of whole-genome sequencing (WGS), has enabled more detailed evaluation of intra-host evolution of viral genomes [12, 13].

Here, we report on the intra-host evolution of B19V during relapsing infection in three immunocompromised patients. We determined genome-wide mutations in the B19V genome from plasma of these patients, and investigate *in silico* the potential conformational changes in the mutated viral proteins using novel machine learning-based modeling. Additionally, we provide a literature overview on intra-host and inter-host B19V genome evolution.

## Methods

### Patients

All patients with a known relapsing or prolonged B19V infection were selected from the laboratory records of the Leiden University Medical Centre (LUMC) in the period 2011-2020. For each patient, four to six B19V DNA-positive serial samples from the laboratory archives were selected for WGS. Samples had previously been sent to the Clinical Microbiology Laboratory (CML) as part of routine patient care for B19V diagnostics, at the discretion of the treating physician. Plasma samples were stored at -80 ×C until WGS analysis.

### Ethics statement

This study has been approved by the Medical Ethics Committee of the LUMC (B20.002, Biobank Infectious Diseases 2020-03 and B20.014, 2020-04).

### B19V PCR and whole genome sequencing

Viral loads were determined by quantitative PCR as previously described (14). Whole genome sequencing was performed using the Arc Bio Galileo Pathogen Solution kit, a complete kit and protocol for quantitative metagenomic detection of several DNA viruses in blood of immunocompromised patients, as previously described (15). In short, patient samples were spiked with an internal baculovirus control before extraction. Nucleic acids were extracted from plasma using the DNA and Viral NA small volume extraction kit on the MagNAPure 96 system (Roche diagnostics, Almere, The Netherlands). After concentration, library preparation was performed with the Galileo Viral Panel sequencing kit (Arc Bio (present: Cantata Bio), LLC, Cambridge, MA, USA) according to the manufacturer’s instructions. Samples were sequenced using the NovaSeq 6000 platform (Illumina, San Diego, CA, USA) at GenomeScan B. V. (Leiden, the Netherlands).

### Bioinformatic analysis

Sequence reads were demultiplexed using bcl2fastq (version 2.2.0) (Illumina, San Diego, CA, USA), resulting in FASTQ files. *De novo* assembly was performed using SPAdes (version 3.11.1). Contigs were mapped against B19V reference genome NC_000883.2. Threshold for nucleotide consensus was set at > 50%. FASTA files were uploaded in Geneious version 2024.0.3 for further comparative and phylogenetic analysis.

### Phylogenetic analysis

All available (near-) complete genome sequences (4800 to 5596 bp) of taxonomy ID 10798 (‘parvovirus B19’) and taxonomy ID 344889 (‘unclassified erythrovirus’) were downloaded from the NCBI database. Clonal and artificial sequences were excluded. Alignments were created with Geneious (version 2024.0.3) A Neighbour-Joining phylogenetic tree (Jukes-Cantor model) was constructed. A second tree was constructed using the NS1-VP1/VP2 section of the genomes (≥4280 bp) in relation to European GenBank genotype 1 submissions.

### Protein structural modeling

Protein sequences with and without the mutations were subjected to protein structure prediction using Alphafold 2 [16]. Mol* was used to visualize the predicted structures and map the predicted local distance difference test (pLDDT) values as heat color on the structure [17]. pLDDT is a per-residue measure of local confidence as predicted by AlphaFold with higher scores indicating a more accurate prediction. In addition to the structure predicted by AlphaFold, the computed pLDDT values reflect the molecular dynamic of the predicted structure [18]. Default parameters in AlphaFold were used after updating all required databases as of June 2023. The evaluation focused on the relaxed models.

## Results

In the period 2011-2020, three patients were identified with relapsing B19V infection. Clinical background, the course of viremia, hemoglobulin levels and immune parameters, including the presence of antibodies and lymphocyte counts, are shown in Figure 1. Patient A and B received a kidney transplantation at t = 0 weeks, at which time both patients tested negative for B19V viremia. Patient C received a stem cell transplantation (SCT) for Non-Hodgkin lymphoma (NHL) 5 months previously, and t = 0 represents the latest time-point before B19V viremia. During most of the sampling period, patients A and B were lymphopenic due to immunosuppressive treatment. While stable donor chimerism was initially achieved after SCT in patient C, lymphocyte function can be considered progressively impaired due to underlying progressive Non-Hodgkin lymphoma (NHL) after SCT from t = 0 onwards. This patient received donor lymphocyte infusions (DLI) as part of NHL treatment during the sample period. Patient A and B were B19V seronegative before transplantation. Patient C was B19V seropositive before transplantation, but the SCT donor was B19V seronegative. Patient A first became IgM-positive, then IgG-positive from 3 weeks after transplantation, but IgG reversion occurred between 20 and 24 weeks after transplantation. Patient B became IgG-positive after IVIG treatment at 12 weeks after transplantation. Patient C first became IgM-positive, then IgG-positive from t = 7 weeks.

**Figure 1:**
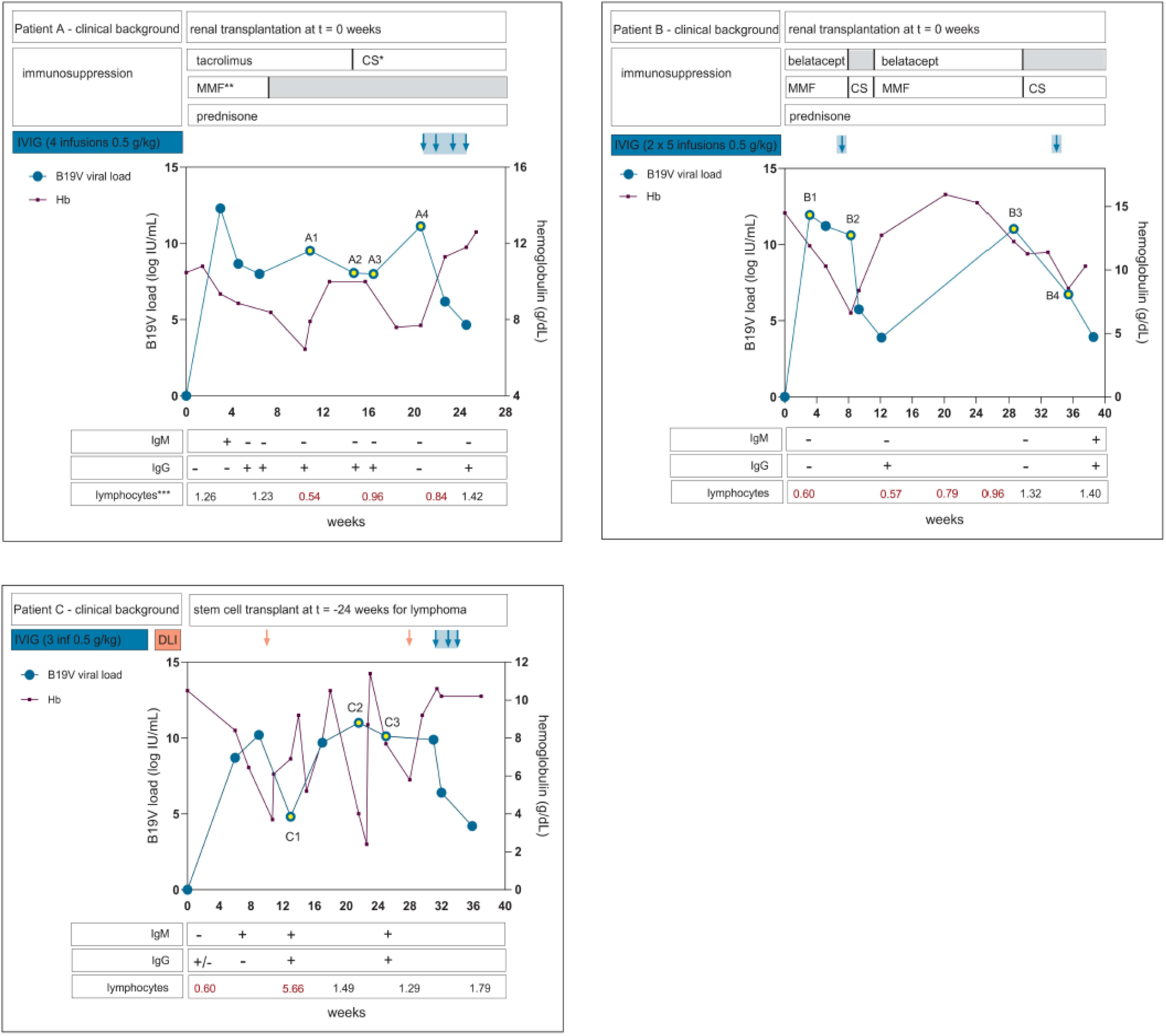
Clinical background, longitudinal course of laboratory parameters and treatment of 3 patients with relapsing B19V infection. *CS = ciclosporin; **MMF = mycophenolate mofetil; ***lymphocytes (x10E9/L), reference values 1-3.5

In patients B and C, viral genomes remained unaltered over a course of 3 months and 6 months of follow-up, respectively: no (non-)synonymous mutations were detected by whole genome sequencing. In patient A, several mutations were identified over a course of 3 months. The first mutation, C3742G, occurred at 14 weeks of follow-up and remained present at 16 weeks of follow-up, while B19V IgG was positive, and was no longer detected in the sample at 20 weeks of follow-up when B19V IgG was negative (Figure 1). At 20 weeks of follow-up, mutation A3982T was detected. From this time point onwards, IVIG was administered and the patient started clearing the infection; no samples with sufficiently high loads were available for sequencing. These mutations were not detected at any time point in patients B and C. Table 1 shows an overview of the detected mutations and the corresponding amino acid changes in patient A.

**Table 1:** overview of nucleotide substitutions and associated amino-acid substitutions from a relapsing infection in patient A.

|  |  | Nt substitution | AA substitution | AA substitution | AA substitution |
| --- | --- | --- | --- | --- | --- |
|  | t |  | VP1 | VP2 | NS |
| Patient A | 0 weeks |  |  |  |  |
|  | 10 weeks | - | - | - | - |
|  | 14 weeks | C3742G | T372S | T145S | na* |
|  | 16 weeks | C3742G | T372S | T145S | na |
|  | 20 weeks | A3892T | Q422L | Q195L | na |

Na = not applicable; no synonymous Nt substitutions were detected in patients A-C (whole genome).

To assess the impact of amino acid changes on the structure of the virus, protein structural modeling was performed on the mutated VP1 and VP2 proteins of B19V as detected in patient A. The C3742G mutation (patient A) translates into a T145S substitution in VP2 which had no predicted impact on its structure. Likewise, the A3892T mutation resulting in a Q422L/Q195L substitution was predicted not to impact VP1/VP2 structure. Notably, the C3742G mutation and associated amino-acid substitution T372S resulted in a predicted conformational change in VP1 – whilst modeling indicated no effect of the same mutation on VP2 conformation. Figure 2 shows the modelled protein structure of the baseline VP1/2 and the modelled protein structure with the amino-acid substitution T372S. The T372S structure shows an additional conformational loop in VP1. The high pLDDT-values resulting in the loop indicate that the T372S structure has changed from a static to a dynamic structure. The predicted additional loop appears adjacent to the primary antibody binding site (VP1u), suggesting that possible reduced antibody binding may be caused by the conformational changes and the resulting steric hindrance.

**Figure 2:**
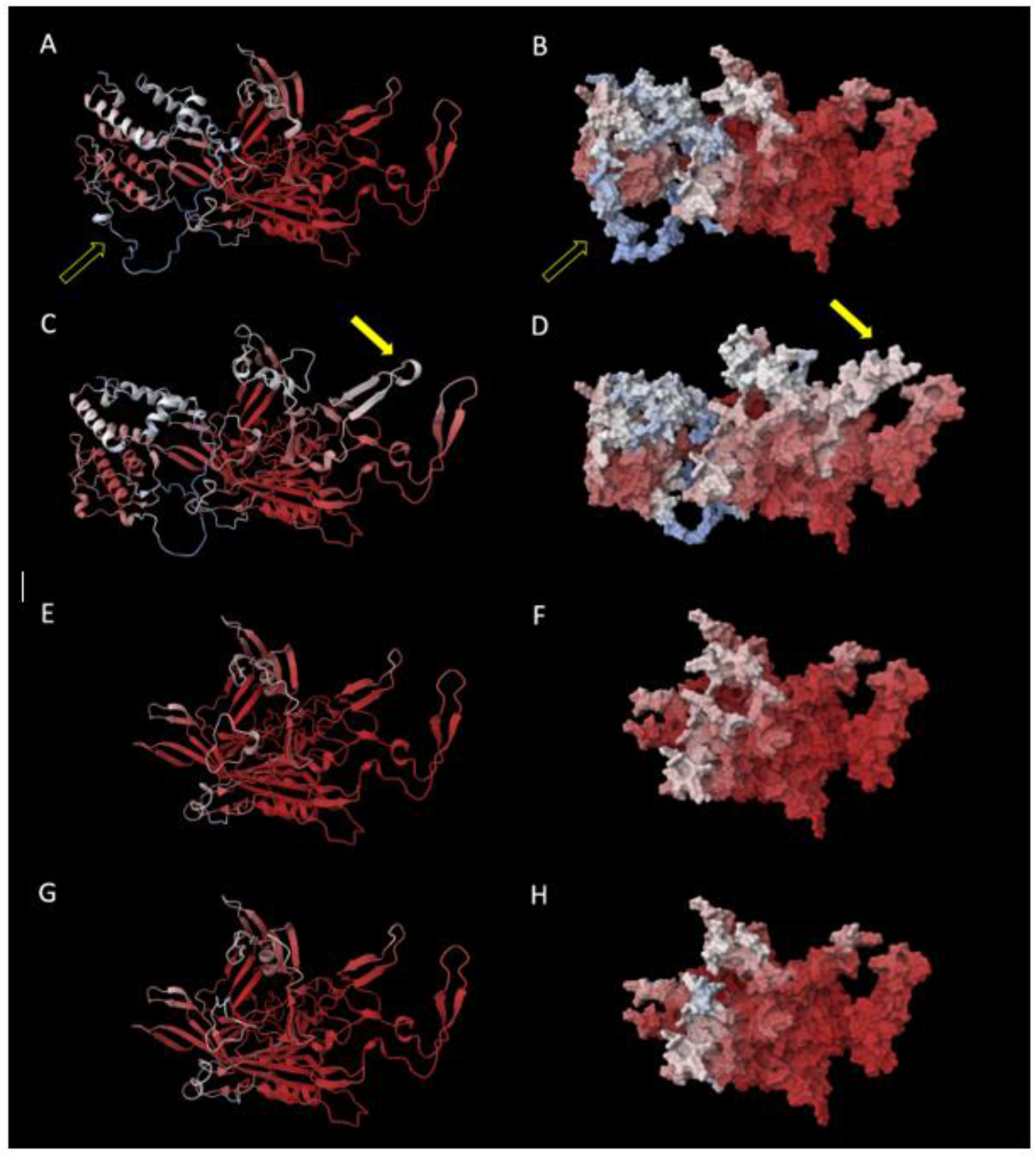
Protein structural modeling of the mutant VP1 and VP2 AA sequence from patient A. 2A: baseline protein structure of VP1 as modelled from the AA sequence for time point A1 (cartoon representation), open arrow indicates VP1u-region (for a view of the entire VP1u-region, see supplementary figure S1); 2B: surface representation of 2A; 2C: protein structure of VP1 as modelled from the AA sequence with T372S substitution (timepoint A2), resulting in an additional loop marked by the closed yellow arrow. 2D: surface representation of 2C; 2E: baseline protein structure of VP2 as modelled from the AA sequence for time point A1 (cartoon representation); 2F: surface representation of 2E; 2G: protein structure of VP2 as modelled from the AA sequence with T372S substitution (timepoint A2), not resulting in structural changes; 2H: surface representation of 2G. The heat color indicate pLDDT values (see methods); red indicates high values.

We also constructed a phylogenetic tree using (near) whole genome sequences from GenBank to assess divergence from other published B19V genotype 1-3 viruses (Figure 3). The viruses from patient A (2019) and B (2015) clustered with genotype 1a, while the virus from patient C (2011) clustered with genotype 3a. The viruses from patient A clustered most closely with relatively recent strains from Serbia (2011) and other recent viruses from France (2017). The viruses from patient B were relatively distant from patient A, but still clustered with genotype 1a. Patient C was infected with genotype 3, which is uncommon in Europe and it is unclear how this patient got infected with a genotype 3 strain.

**Figure 3:**
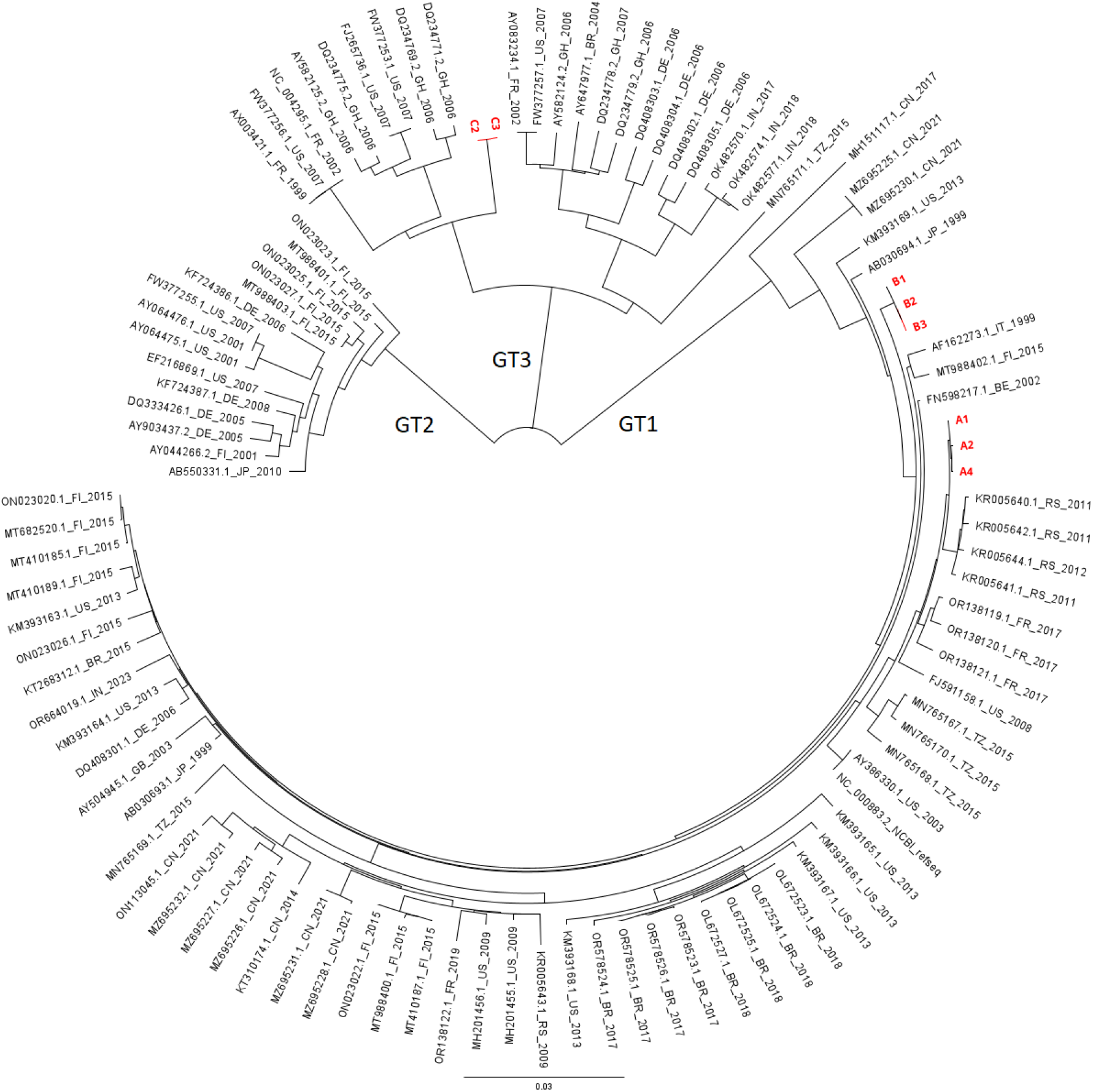
Phylogenetic analysis of patient’s A and B sequences and global B19V GenBank strains. Selected Genbank strains > 4800 bp of genotype 1, 2 and 3. GenBank strains are denoted by GenBank entries followed by country (2-digit code according to ISO-3166-1 alpha 2 country codes) and year of isolation.

To assess the relation to other local strains, a separate tree was constructed using European GenBank sequences, analyzing a subregion of the genome (4280 bp) which includes the complete NS1-VP1/2 fragments. The viruses from patient A and B clustered closely with other national strains within genotype 1a (supplemental figure S1). As there were very few European GenBank entries for genotype 3, patient C is not included in this analysis.

## Discussion

In this study, we investigated the intra-host evolution of B19V in immunocompromised patients with relapsing B19V infection. In two patients, the viral genome remained stable over several months, despite multiple episodes of intense replication and treatment with IVIG during the sample period in one of the patients. In one patient, two non-synonymous/missense mutations were detected in the VP1/VP2 region, resulting in AA substitutions in 4 locations. These mutations have not previously been described in the literature (table 1) and were not present in GenBank sequences used in our study. Modeling of protein folding predicted that one of these mutations (C3742G) would result in a conformational change in VP1.

The B19V capsid is composed of VP1 (5%) and VP2 (95%), while the NS protein is not expressed on the surface. Therefore, VP1 and VP2 are considered the most antigenic, which has also been shown in immunization assays [19, 20]. The main antigenic domain is thought to be the ‘VP1 unique region’ (VP1u), a protein structure consisting of 227 amino acids on the B19V capsid involved in receptor binding (see figure 2 and supplemental figure S1) [21]. We hypothesized that humoral immune pressure and specifically IVIG might be drivers for B19V evolution and we expected that changes would most likely occur in the VP1u-region. Although we did observe mutations under humoral immune pressure, these mutations did not occur in the region coding for VP1u. Also, modeling of protein folding predicted a conformational change in VP1 but not at the site of VP1u (figure 2).

However, the conformational change indicates a change from a static to a dynamic structure, and it is not yet clear how this may affect the interaction with surrounding molecules. To assess the possible role of the predicted conformational change in humoral immune evasion, its position in relation to the complete capsid structure should be evaluated. Future research should therefore firstly focus on changes in capsid structure. Subsequently, molecular docking studies, in depth molecular dynamic simulations and in vitro binding studies would be needed to understand the impact of the predicted molecular changes on e.g. antibody binding.

In patient A the first mutations emerged when the patient was B19V IgG-seropositive. The serological profile of patient A indicated a pattern most compatible with natural immunity, the appearance (and disappearance) of IgM followed by prolonged detection of strong-positive signals of IgG. Remarkably, IgG-seroreversion occurred shortly after the appearance of mutations (fig 1). The patient had not yet received IVIG or other blood products that may explain transient IgG-positivity, so most likely this indicates a loss of natural immunity, which may be considered rare. In patient B, who was sampled during IVIG treatment, no mutations occurred. Patient C was not sampled during IVIG-treatment but did receive DLI during the sampling period. This might have resulted in a decrease in viral load (although the SCT donor was B19V seronegative) (Figure 1). Notably, this primarily T-cell mediated treatment did apparently not lead to selection of variants. These results suggest that natural humoral immune pressure might play a more important role in intra-host evolution than IVIG and T-cell-mediated immunity.

Previous studies on B19V intra-host evolution have shown varying results. Table 2 shows an overview of available literature on B19V intra-host evolution [8-11, 22]. One study on intra-host evolution found a relatively high mutation rate compared to other studies [9]. In this study by Hung et al, B19V longstanding infection was studied in three AIDS patients and a large number of single nucleotide polymorphisms (SNPs) were observed during a maximum follow-up of 11 months. Remarkably, the large number of mutations only occurred in the two patients treated with HAART. Only one mutation was found to appear under IVIG treatment. Of note, none of the new strains became dominant. In our study, we observed a disappearance of the nonsynonymous substitution after seroreversion at 18 weeks of follow-up. Combined with the other studies in which relatively few changes in the viral genome were detected in long-term infection and reversion of these changes in the course of follow-up, these studies suggest a relatively high level of genetic stability. As we only found mutations occurring while natural immunity was developing, it could be hypothesized that humoral immune selection is stronger with natural immunity than with IVIG treatment. As we investigated a limited patient population, further studies with larger patient populations would be required to investigate this hypothesis.

**Table 2:** literature overview of studies on intra-host evolution of B19V.

| First author | year | Study population | B19V treatment | Sequencing method | Nucleotide region sequenced | Follow-up period | results |
| --- | --- | --- | --- | --- | --- | --- | --- |
| Gallinella | 1996 | 1 patient; chronic anemia | Not mentioned | Sanger | 2400-3400 | 16 months | No changes in viral genome |
| Plentz | 2004 | 1 patient; bone marrow transplantation | IVIG | Not mentioned | Not mentioned | 8 months | 3 lasting changes after temporary variations:<br>T3463C<br>C4852G<br>T4867C |
| Hung | 2006 | 3 patients; AIDS | IVIG (n=3); HAART (n=2) | Sanger | 436 – 2431;<br>3125- 4283 | 11 months | Pt 1: A3271C<br>Pt 2: 18 SNP (14 N*)<br>Pt 3: 15 SNP (9 N) |
| Suzuki | 2014 | 1 pediatric patient; cord blood transplantation | IVIG | Sanger | 602-5014 | 29 months | 6 SNP<br>T/C941T<br>T/C1037T<br>A1048A/G (N)<br>T1112C<br>T1118T/C<br>A1266A/G (N) |
| Jain | 2018 | 13 pediatric hematological malignancy patients followed up; 3 patients with at least one mutation detected | Not mentioned | Sanger | 1747-2691 | 6 months | Pt 1: G579A<br>Pt 2: C577G<br>Pt 3: C672G |

\*N = nonsynonymous mutation

Whole-genome phylogenetic analysis on the viruses of our patients showed that our viral sequences clustered with other circulating viral strains, suggesting these are representative strains for these regions and related to other Dutch and European strains. Studies on population evolution fairly consistently report a substitution rate of ∼1 × 10^−4^ substitutions per site per year (s/s/y) for B19V, with highest s/s/y reported for the capsid sequences VP1 and VP2 (for an overview of studies, see supplementary table 1) (8, 23-28, 31). This is a relatively high substitution rate for DNA viruses, as substitution rates for other DNA viruses are estimated at 10^−5^ to 10^−9^ s/s/y, although with higher s/s/y for single-stranded DNA viruses (such as B19V) [29]. However, these studies on B19V are based on a relatively short period of observance which may cause an overestimation of mutation rate [30]. From an evolutionary point-of-view, B19V intra-host genomic stability is plausible considering the long-term history that B19V and mankind share, as B19V was already demonstrated in 7000-year old human bone samples [27]. In these studies investigating ancient DNA, substitution rates between 1.02-1.22 × 10^−5^ s/s/y were reported [27, 31].

Despite the observed relative genetic stability on population level, much remains unknown on intra-host viral evolution. B19V relapsing infections are a relatively new phenomenon. It is only in the last decades that the circumstances in which B19V may relapse have become present; the development of transplantation medicine has created the existence of consistently severely immunocompromised hosts that may form a reservoir for viral persistence and evolution. In addition, specific treatments such as IVIG may still add to selection pressure and data on intra-host evolution are still scarce for B19V. We recommend to monitor genome changes and their structural impact on the virus in larger series to increase our understanding of viral evolution under these relatively new circumstances.

## Data Availability

All data produced in the present study are available upon reasonable request to the authors

## Acknowledgments

We thank Ellen Carbo for technical assistance in sequencing and Roel Nijhuis for assistance with the use of Geneious software.

## Conflict of interest

none reported

## Supplementary information

**Figure S1:**
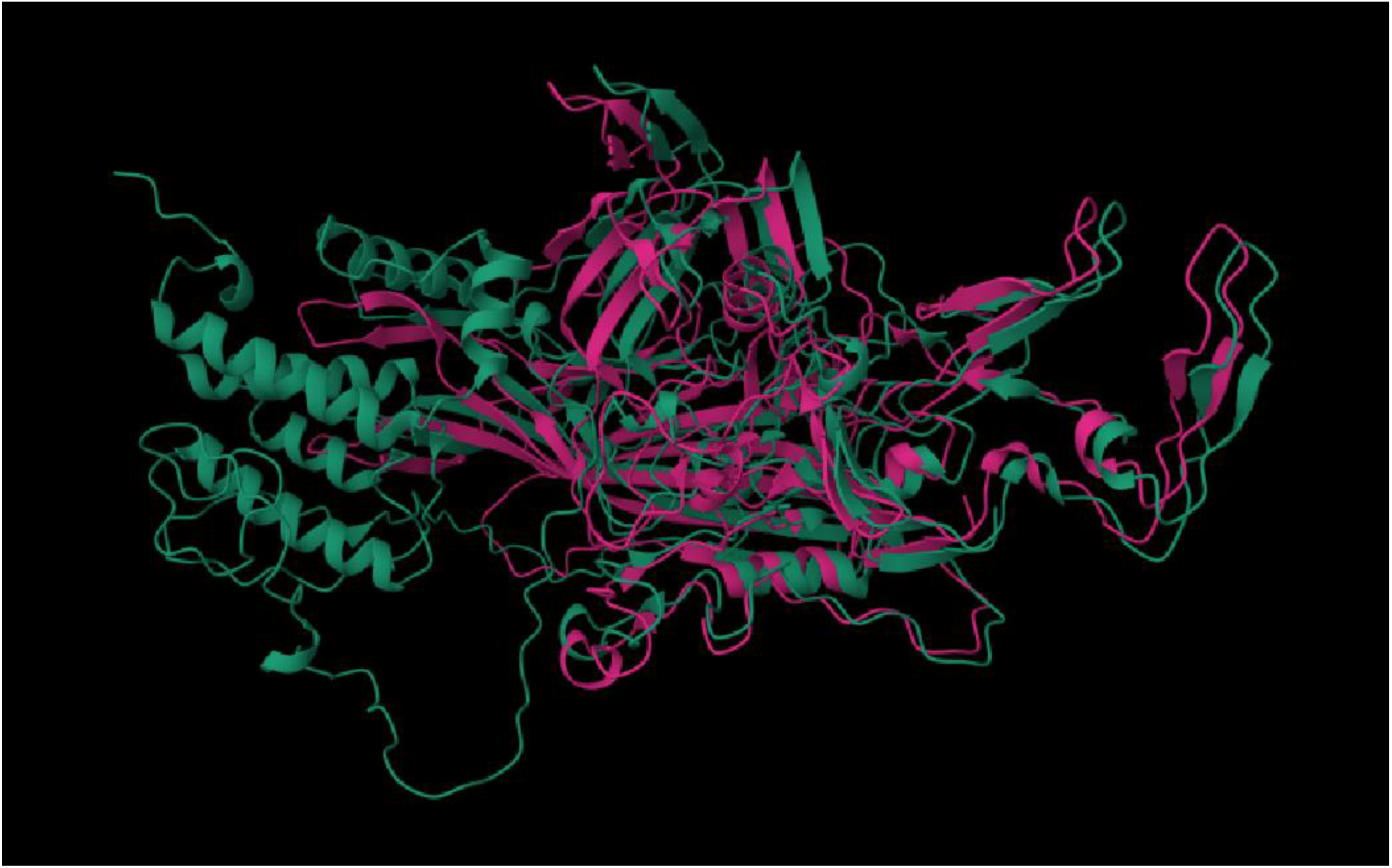
Protein structural deep learning modeling of VP1 (green) and VP2 (pink) in a superimposed view. The area of VP1 that does not overlap with VP2 is the VP1-unique region, which holds the receptor-binding domain.

**Figure S2:**
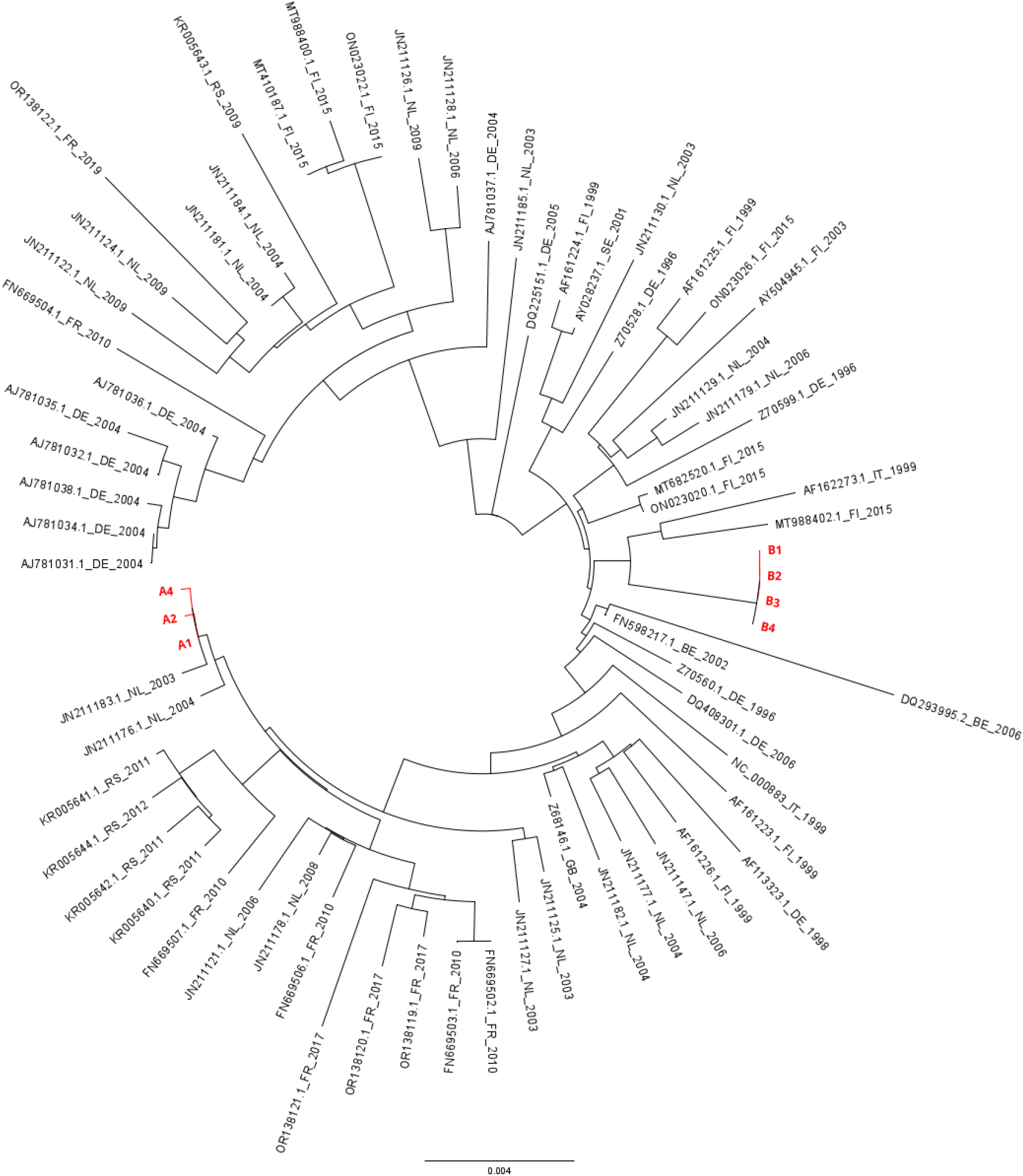
Phylogenetic analysis of NS1,VP1/2 (4280 bp) of study isolates in relation to other European genotype 1 strains. GenBank strains are denoted by GenBank entries followed by country (2-digit code according to ISO-3166-1 alpha 2 country codes) and year of isolation.

**Supplementary table 1:** literature overview of studies on population evolution of B19V.

| First author | year | Study population | Geographical sampling location | Sampling period | Sequencing method | Results |
| --- | --- | --- | --- | --- | --- | --- |
| Gallinella | 1996 | Blood samples from 7 known B19V patients; nt 2400-3400 (VP) sequenced | Italy | 1989-1994 | Sanger | No s/s/y* reported; divergence rate within geographic cluster similar to global divergence |
| Shackelton | 2006 | Data set B19V genotype 1: 38 VP1 sequences; 27 NS1-VP1 sequences | Europe (England, Germany, Finland, Ireland, Sweden); USA; Brazil; Japan; Korea; China | 1973-2001 | Gen Bank sequences | Both VP1 and NS1: $\sim 1 \times 10^{-4}$ nucleotide s/s/y |
| Norja | 2008 | 63 blood or autopsy samples; Nt 3242-4612 (VP) sequenced | Europe (Germany, Finland, UK) | 1980-2007 | Sanger | $4 \times 10^{-4}$ s/s/y |
| Suzuki | 2008 | 104 NS1-VP1 junction sequences | Sapporo (Japan) | 1980-2008 | Sanger | No s/s/y reported; 2 distinct patterns observed in small region: 1. accumulation of SNP and 2. Sudden replacement of strains |
| Toppinen | 2015 | 43 bone samples; part of NS1/VP sequenced | Finland | WWII casualties | Sanger (?) | $1.1-3.2 \times 10^{-4}$ s/s/y |
| Stamenkovic | 2016 | 137 Gen Bank sequences nt 665-4851 | Europe; USA; Brazil; Japan; Vietnam | 1973-2012 | Gen Bank sequences | $1.03 \times 10^{-4}$ s/s/y |
| Mühlemann | 2018 | 10 samples of preserved ancient B19V from teeth (63.9 - 99.7% coverage) | Eurasia; South-East Asia; Greenland | $\sim 4800$ BC – 1000 AD | NGS | $1.22 \times 10^{-5}$ s/s/year (strict clock) |
| Guzman-Solís | 2021 | 3 samples of preserved ancient B19V from teeth (92.4-99.1% coverage) | Mexico | 16<sup>th</sup>- 18<sup>th</sup> century | NGS | $1.03 \times 10^{-5}$ s/s/y (strict clock) |

\*s/s/y = substitutions/site/year^; ^bone material from WWII casualty graves

## Notes

### Competing Interest Statement

The authors have declared no competing interest.

### Funding Statement

This study did not receive any funding

## References

1. Ganaie SS, Qiu J. Recent Advances in Replication and Infection of Human Parvovirus B19. Front Cell Infect Microbiol. 2018;8:166.

2. Eid AJ, Ardura MI. Human parvovirus B19 in solid organ transplantation: Guidelines from the American society of transplantation infectious diseases community of practice. Clin Transplant. 2019;33(9):e13535.

3. Rosado-Canto R, Carrillo-Pérez DL, Jiménez JV, Cuellar-Rodríguez J, Parra-Avila I, Alberú J, et al. Treatment strategies and outcome of parvovirus B19 infection in kidney transplant recipients: a case series and literature review of 128 patients. Rev Invest Clin. 2019;71(4):265–74.

4. Oude Munnink BB, Worp N, Nieuwenhuijse DF, Sikkema RS, Haagmans B, Fouchier RAM, et al. The next phase of SARS-CoV-2 surveillance: real-time molecular epidemiology. Nat Med. 2021;27(9):1518–24.

5. Gooskens J, Jonges M, Claas EC, Meijer A, Kroes AC. Prolonged influenza virus infection during lymphocytopenia and frequent detection of drug-resistant viruses. The Journal of infectious diseases. 2009;199(10):1435–41.

6. Campos AB, Ribeiro J, Boutolleau D, Sousa H. Human cytomegalovirus antiviral drug resistance in hematopoietic stem cell transplantation: current state of the art. Reviews in medical virology. 2016;26(3):161–82.

7. Munnink BBO, Nijhuis RHT, Worp N, Boter M, Weller B, Verstrepen BE, et al. Highly Divergent SARS-CoV-2 Alpha Variant in Chronically Infected Immunocompromised Person. Emerg Infect Dis. 2022;28(9):1920–3.

8. Gallinella G, Venturoli S, Gentilomi G, Musiani M, Zerbini M. Extent of sequence variability in a genomic region coding for capsid proteins of B19 parvovirus. Arch Virol. 1995;140(6):1119–25.

9. Hung CC, Sheng WH, Lee KL, Yang SJ, Chen MY. Genetic drift of parvovirus B19 is found in AIDS patients with persistent B19 infection. Journal of medical virology. 2006;78(11):1374–84.

10. Suzuki M, Ito Y, Shimada A, Saito M, Muramatsu H, Hama A, et al. Long-term parvovirus B19 infections with genetic drift after cord blood transplantation complicated by persistent CD4+ lymphocytopenia. J Pediatr Hematol Oncol. 2014;36(1):e65–8.

11. Plentz A, Hahn J, Holler E, Jilg W, Modrow S. Long-term parvovirus B19 viraemia associated with pure red cell aplasia after allogeneic bone marrow transplantation. Journal of clinical virology : the official publication of the Pan American Society for Clinical Virology. 2004;31(1):16–9.

12. Lu IN, Muller CP, He FQ. Applying next-generation sequencing to unravel the mutational landscape in viral quasispecies. Virus Res. 2020;283:197963.

13. Huang SW, Hung SJ, Wang JR. Application of deep sequencing methods for inferring viral population diversity. J Virol Methods. 2019;266:95–102.

14. Knoester M, von dem Borne PA, Vossen AC, Kroes AC, Claas EC. Human parvovirus B19 genotype 3 associated with chronic anemia after stem cell transplantation, missed by routine PCR testing. Journal of clinical virology : the official publication of the Pan American Society for Clinical Virology. 2012;54(4):368–70.

15. Carbo EC, Russcher A, Kraakman MEM, de Brouwer CS, Sidorov IA, Feltkamp MCW, et al. Longitudinal Monitoring of DNA Viral Loads in Transplant Patients Using Quantitative Metagenomic Next-Generation Sequencing. Pathogens. 2022;11(2).

16. Tunyasuvunakool K, Adler J, Wu Z, Green T, Zielinski M, Žídek A, et al. Highly accurate protein structure prediction for the human proteome. Nature. 2021;596(7873):590–6.

17. Sehnal D, Bittrich S, Deshpande M, Svobodová R, Berka K, Bazgier V, et al. Mol* Viewer: modern web app for 3D visualization and analysis of large biomolecular structures. Nucleic Acids Res. 2021;49(W1):W431–w7.

18. Guo HB, Huntington B, Perminov A, Smith K, Hastings N, Dennis P, et al. AlphaFold2 modeling and molecular dynamics simulations of an intrinsically disordered protein. PLoS One. 2024;19(5):e0301866.

19. Saikawa T, Anderson S, Momoeda M, Kajigaya S, Young NS. Neutralizing linear epitopes of B19 parvovirus cluster in the VP1 unique and VP1-VP2 junction regions. Journal of virology. 1993;67(6):3004–9.

20. Anderson S, Momoeda M, Kawase M, Kajigaya S, Young NS. Peptides derived from the unique region of B19 parvovirus minor capsid protein elicit neutralizing antibodies in rabbits. Virology. 1995;206(1):626–32.

21. Ros C, Bieri J, Leisi R. The VP1u of Human Parvovirus B19: A Multifunctional Capsid Protein with Biotechnological Applications. Viruses. 2020;12(12).

22. Jain A, Jain P, Kumar A, Prakash S, Khan DN, Kant R. Incidence and progression of Parvovirus B19 infection and molecular changes in circulating B19V strains in children with haematological malignancy: A follow up study. Infect Genet Evol. 2018;57:177–84.

23. Shackelton LA, Holmes EC. Phylogenetic evidence for the rapid evolution of human B19 erythrovirus. Journal of virology. 2006;80(7):3666–9.

24. Suzuki M, Yoto Y, Ishikawa A, Tsutsumi H. Analysis of nucleotide sequences of human parvovirus B19 genome reveals two different modes of evolution, a gradual alteration and a sudden replacement: a retrospective study in Sapporo, Japan, from 1980 to 2008. Journal of virology. 2009;83(21):10975–80.

25. Toppinen M, Perdomo MF, Palo JU, Simmonds P, Lycett SJ, Soderlund-Venermo M, et al. Bones hold the key to DNA virus history and epidemiology. Sci Rep. 2015;5:17226.

26. Stamenković GG, Ćirković VS, Šiljić MM, Blagojević JV, Knežević AM, Joksić ID, et al. Substitution rate and natural selection in parvovirus B19. Sci Rep. 2016;6:35759.

27. Mühlemann B, Margaryan A, Damgaard PB, Allentoft ME, Vinner L, Hansen AJ, et al. Ancient human parvovirus B19 in Eurasia reveals its long-term association with humans. Proceedings of the National Academy of Sciences of the United States of America. 2018;115(29):7557–62.

28. Norja P, Hokynar K, Aaltonen LM, Chen R, Ranki A, Partio EK, et al. Bioportfolio: lifelong persistence of variant and prototypic erythrovirus DNA genomes in human tissue. Proceedings of the National Academy of Sciences of the United States of America. 2006;103(19):7450–3.

29. Duffy S, Shackelton LA, Holmes EC. Rates of evolutionary change in viruses: patterns and determinants. Nat Rev Genet. 2008;9(4):267–76.

30. dos Reis M, Donoghue PC, Yang Z. Bayesian molecular clock dating of species divergences in the genomics era. Nat Rev Genet. 2016;17(2):71–80.

31. Guzmán-Solís AA, Villa-Islas V, Bravo-López MJ, Sandoval-Velasco M, Wesp JK, Gómez-Valdés JA, et al. Ancient viral genomes reveal introduction of human pathogenic viruses into Mexico during the transatlantic slave trade. Elife. 2021;10.

